# Estimating healthcare resource needs for COVID-19 patients in Nigeria

**DOI:** 10.1101/2020.08.19.20178434

**Authors:** Adeteju Ogunbameru, Kali Barrett MD, Arinola E. Joda, Yasin A Khan MD, Petros Pechlivanoglou, Stephen Mac, David Naimark MD, Raphel Ximenes, Beate Sander

## Abstract

**Background:** Predicting potential healthcare resource use under different scenarios will help to prepare the healthcare system for a surge in COVID-19 patients. In this study, we aim to predict the effect of COVID-19 on hospital resources in Nigeria.

**Method:** We adopted a previously published discrete-time, individual-level, health-state transition model of symptomatic COVID-19 patients to the Nigerian healthcare system and COVID-19 epidemiology. We simulated different combined scenarios of epidemic trajectories and acute care capacity. Primary outcomes included expected cumulative number of cases, days until depletion resources, and number of deaths associated with resource constraints. Outcomes were predicted over a 60-day time horizon.

**Results:** In our best-case epidemic trajectory, which implies successful implementation of public health measures to control COVID-19 spread, the current number of ventilator resources in Nigeria (conservative resources scenario), were expended within five days, and 901 patients may die while waiting for hospital resources in conservative resource scenario. In our expanded resource scenarios, ventilated ICU beds were depleted in all three epidemic trajectories within 60 days. Acute care resources were only sufficient in the best-case and intermediate epidemic scenarios, combined with a substantial increase in healthcare resources.

**Conclusion:** Current hospital resources are inadequate to manage the COVID-19 pandemic in Nigeria. Given Nigeria’s limited resources, it is imperative to increase healthcare resources and maintain aggressive public health measures to reduce COVID-19 transmission.

**KEY QUESTIONS:** *What is already known on this subject?:* While western countries seem to be recovering from the COVID-19 pandemic, there is an increasing community spread of the virus in many African countries. The limited healthcare resources available in the region may not be sufficient to cope with increasing numbers of COVID-19 cases.

*What this study adds?:* Using the COVID-19 Resource Estimator (CORE) model, we demonstrate that implementing and maintaining aggressive public health measures to keep the epidemic growth at a low rate, while simultaneously substantially increasing healthcare resources is critical to minimize the impact of COVID-19 on morbidity and mortality. The impact of COVID-19 in low resource settings will likely overwhelm health system capacity if aggressive public health measures are not implemented. To mitigate the impact of COVID-19 in these settings, it is essential to develop strategies to substantially increase health system capacities, including hospital resources, personal protective equipment and trained healthcare personnel and to implement and maintain aggressive public health measures.

## INTRODUCTION

The World Health Organization declared the spread of severe acute respiratory syndrome coronavirus 2 (SARS-CoV-2) a Public Health Emergency of International concern on January 30, 2020, and later a pandemic on March 11, 2020.^1^ Africa recorded the first case of Coronavirus Disease 2019 (COVID-19) on February 14, 2020, in Egypt^2^, and since then has reported over 945,165 cases and 18,476deaths as of August 17, 2020.^3^

Nigeria reported the first case of COVID-19 on February 29, 2020.^4^ By August 12, 47,743confirmed cumulative cases were reported.^4^ The average daily growth rate of cumulative cases in Nigeria from when 100 cases were reported on March 29, 2020 (which correspond to the date lockdown was initiated in the three most affected states) to August 12, 2020 is 5%.^4^ The lockdown ban was lifted on May 4, 2020. The mean growth rate from May 5, 2020 to August 16,2020 is 3%.^4^ From June 1, 2020, to August 12, 2020, the mean growth rate furthered decrease to 2%.^4^ The growth rate of COVID-19 in Nigeria is low compared to what was observed from March to May in Europe and North America.^3^ Overall, the pandemic trajectory in Africa has been different compared to China, Europe and North America, with lower growth rates and lower case fatality rates.^5^ However, the spread of COVID-19 in the region is rapidly increasing, with a shift from 9 to 42 countries affected from March to May 2020.^5^ Factors that have been associated with the rapid spread of COVID-19 virus in the region include poor public health infrastructure, underreporting, and limited testing and contract tracing resources.^6^ To mitigate the impact of COVID-19, aggressive public health measures must be pursued to prevent overwhelming the healthcare system which has limited resources.^7^

To reduce the spread of COVID-19 in Nigeria, the Federal government initiated a Presidential Task Force to provide a high-level strategic national response to the disease; launched a campaign themed “Take Responsibility” – a call to encourage residents to take individual and collective responsibility of implementing public health measures; quarantined all COVID-19 confirmed cases, test and track confirmed cases contacts; banned non-essential international and inter-state travel; initiated curfews to limit social interactions and restricted gathering to 20 people per workplace to encourage physical distancing.^4^

Predicting COVID-19 population spread and healthcare resource needs for symptomatic patients using models is essential to prepare the health system and allow it to continue running efficiently during the pandemic, minimize morbidity, and reduce society disruption.^8^ Simulation models can help to inform healthcare resource needs under different scenarios to improve planning and support procurement strategies, especially in resource-limited settings.^9^

Nigeria, like many African countries, has limited healthcare resources and infrastructure. Before the start of the pandemic, Nigeria had an estimated 0.2 hospital beds per 1,000 population, 350 intensive care-unit (ICU) beds without ventilators (equivalent to 0.07 ICU beds per 100,000 population), and 450 ventilated beds.^10,11,12^ With the country’s cumulative COVID-19 cases exceeding 47,000 on August 11 2020,^4^ and case numbers still increasing, forecasting how COVID-19 will affect hospital resources under different scenarios is critical for Nigeria’s COVID-19 response.

Our objective is to predict the short-term effect of COVID-19 on hospital resources in Nigeria for a range of COVID-19 epidemic and hospital capacity scenarios using a health system simulation model.

## METHODS

### Study design

We adopted the previously published COVID-19 Resource Estimator (CORE) model from Ontario, Canada,^13^ to fit the healthcare system and COVID-19 epidemiology in Nigeria. Our primary outcomes included projected cumulative number of COVID-19 cases, number of days until depletion of ward bed and ventilated ICU bed resources and number of avoidable deaths assuming no resource constraint. Outcomes were predicted over a 60-day time horizon in daily time steps.

### CORE model structure

We used the CORE model that was available as an interactive software application online.^11^ A detailed description of the CORE model is provided elsewhere.^9^ Briefly, CORE is a discrete-time, individual-level, health-state transition model of symptomatic COVID-19 patients. CORE simulates a dynamic population of symptomatic COVID-19 adult patients (18 years and above) who present to the emergency department (ED), where they are either sent home to self-isolate or admitted to the hospital. If admitted, COVID-19 patients are assigned to either a general ward or ICU, depending on disease severity. Seventy-eight percent of ICU patients were assumed to require invasive mechanical ventilation.^9,10^

### CORE model assumptions

Several assumptions were made in the CORE model. For simulated patients in the model, resource needs were based on the patient’s health state (i.e. in a general ward or ICU). If there was a lack of ward or ICU beds, patients remained with their current available resources until the needed resource became available. Patients requiring mechanical ventilation but did not have access were assumed dead within 24 hours. Mortality risk was attributed to only ICU patients with or without a need for a ventilator. Patients awaiting an ICU bed (but not mechanical ventilation) were assumed to have the same risk of death as patients in ICU. Ward beds were assumed available upon patients’ recovery. ICU beds with/without ventilators were assumed to be freed up upon recovery or death of patients. Available resources were prioritized based on the patient’s location (e.g. ward beds will be prioritized to ICU patients before new admissions) and the length of waiting time of a patient since admission. The CORE model assumed that other essential resources, including personal protective equipment and essential medications, would be available in sufficient quantities. It also assumed all available beds are adequately staffed, and no staffing shortage would emerge with vast-increase in hospital resources.

### Assumptions specific to Nigeria hospital setting

We assumed that COVID-19 patients would only be treated in hospitals licensed by the Nigeria Centre for Disease Control.^14^ Patients were assumed to be admitted to ICU only when in need of mechanical ventilation and that all ICU beds are vented.

### Model parameters

All model parameters are listed in Table 1.

**Table 1:**
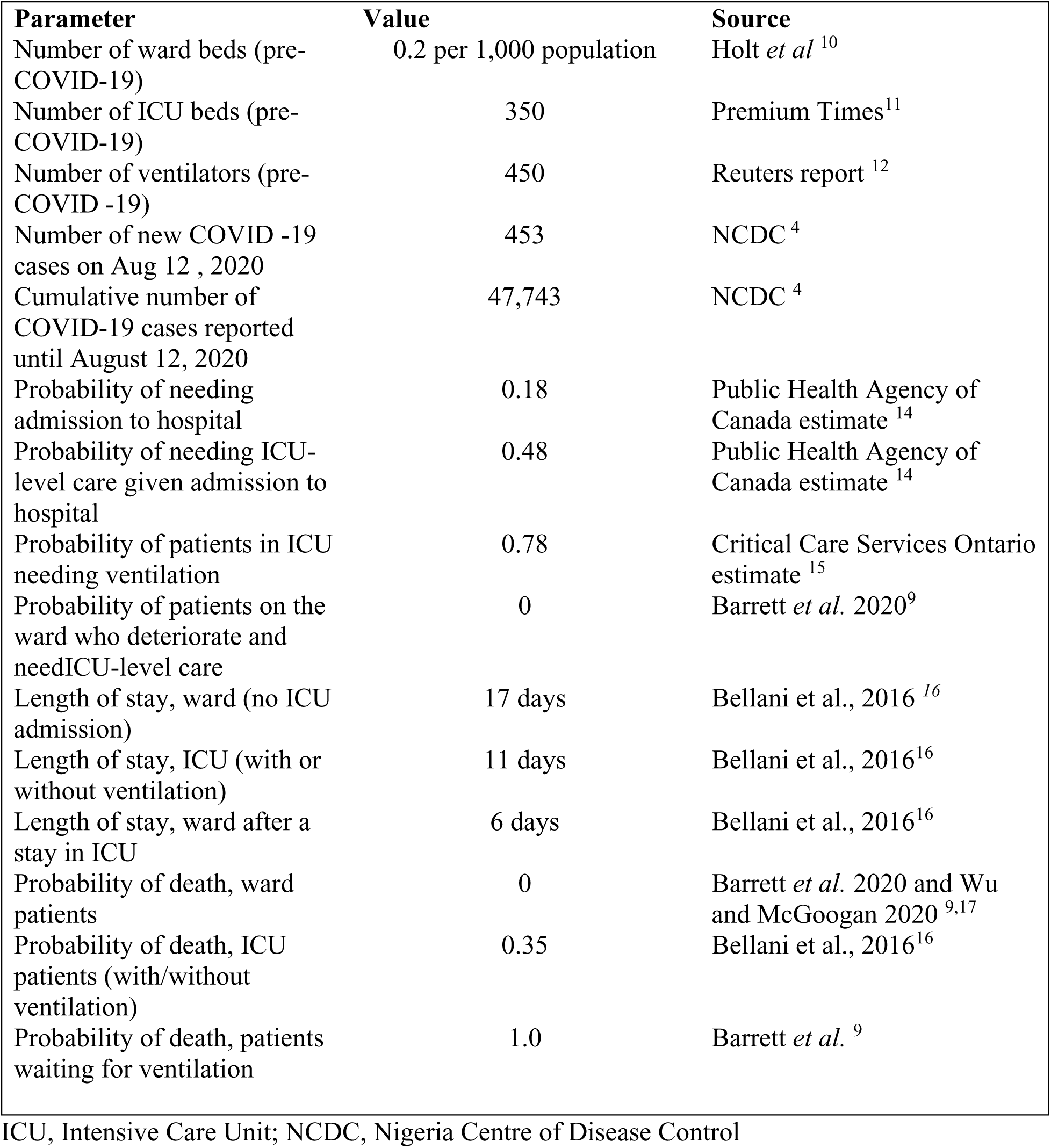
Model parameters Parameter.

| **Parameter** | **Value** | **Source** |
| --- | --- | --- |
| Number of ward beds (pre-COVID-19) | 0.2 per 1,000 population | Holt *et al* <sup>10</sup> |
| Number of ICU beds (pre-COVID-19) | 350 | Premium Times<sup>11</sup> |
| Number of ventilators (pre-COVID -19) | 450 | Reuters report <sup>12</sup> |
| Number of new COVID -19 cases on Aug 12 , 2020 | 453 | NCDC <sup>4</sup> |
| Cumulative number of COVID-19 cases reported until August 12, 2020 | 47,743 | NCDC <sup>4</sup> |
| Probability of needing admission to hospital | 0.18 | Public Health Agency of Canada estimate <sup>14</sup> |
| Probability of needing ICU-level care given admission to hospital | 0.48 | Public Health Agency of Canada estimate <sup>14</sup> |
| Probability of patients in ICU needing ventilation | 0.78 | Critical Care Services Ontario estimate <sup>15</sup> |
| Probability of patients on the ward who deteriorate and need ICU-level care | 0 | Barrett *et al.* 2020<sup>9</sup> |
| Length of stay, ward (no ICU admission) | 17 days | Bellani et al., 2016 <sup>16</sup> |
| Length of stay, ICU (with or without ventilation) | 11 days | Bellani et al., 2016<sup>16</sup> |
| Length of stay, ward after a stay in ICU | 6 days | Bellani et al., 2016<sup>16</sup> |
| Probability of death, ward patients | 0 | Barrett *et al.* 2020 and Wu and McGoogan 2020 <sup>9,17</sup> |
| Probability of death, ICU patients (with/without ventilation) | 0.35 | Bellani et al., 2016<sup>16</sup> |
| Probability of death, patients waiting for ventilation | 1.0 | Barrett *et al.* <sup>9</sup> |

ICU, Intensive Care Unit; NCDC, Nigeria Centre of Disease Control

#### COVID-19 acute resource utilization

The fixed parameters in the CORE model included probabilities of hospital and ICU admission obtained from reported Canadian COVID-19 data, reflecting clinical need without resource constraints.^9,15^ Mean length of hospitalization, and the probability of death for COVID-19 patients in the ward and ICU were based on reported data for patients admitted for moderate acute respiratory distress syndrome (ARDS), based on a similar in clinical manifestation.^9,16,17,18^ The proportion of ICU patients requiring mechanical ventilation was obtained from reported Canadian data.^9,16^

#### Epidemic trajectories

We used reported data on daily and cumulative COVID-19 cases up to August 12, 2020, when the number of cumulative cases exceeded 49,000 as published by the Nigeria Centre for Disease Control (NCDC).^4^ We forecasted three scenarios – best case, intermediate case, and worst case – to predict the trajectory of COVID-19 cases in Nigeria for 60 days starting with data from August 12, 2020. In the three scenarios, we started our prediction with 47,743 confirmed COVID-19 cumulative cases and 453 new cases within 24 hours. For our best-case scenario, we assumed an infection growth rate of 1%, to depict the successful implementation of aggressive public health measures (i.e., social distancing, school closure and travel restrictions). To our knowledge, no African country has an average infection growth rate of 1%, from the day when 100 cases were reported. In our intermediate scenario analysis, we used a 2% infection growth rate, which is the average growth rate from June 1, 2020 to August 12, 2020 in Nigeria.^4^ Similarly, Ghana recorded an average growth of 2% from June 1, 2020 to August 12, 2020.^19^ For our worst-case scenario, we assumed an infection growth rate of 3%, which suggests a scenario where no further public health measures are introduced, economy re-opening is fast-tracked and greater community transmission is ongoing. Infection growth rates across countries were calculated using daily reported COVID-19 cases from the John Hopkins University repository.^19^ We did not assume an epidemic peak in our study since our time horizon is short (60 days), and the daily number of new cases still fluctuates between low and high numbers. Case predictions for the three scenarios are shown in Figure 2 (daily number of cases are provided in Appendix 1).

**Figure 1:**
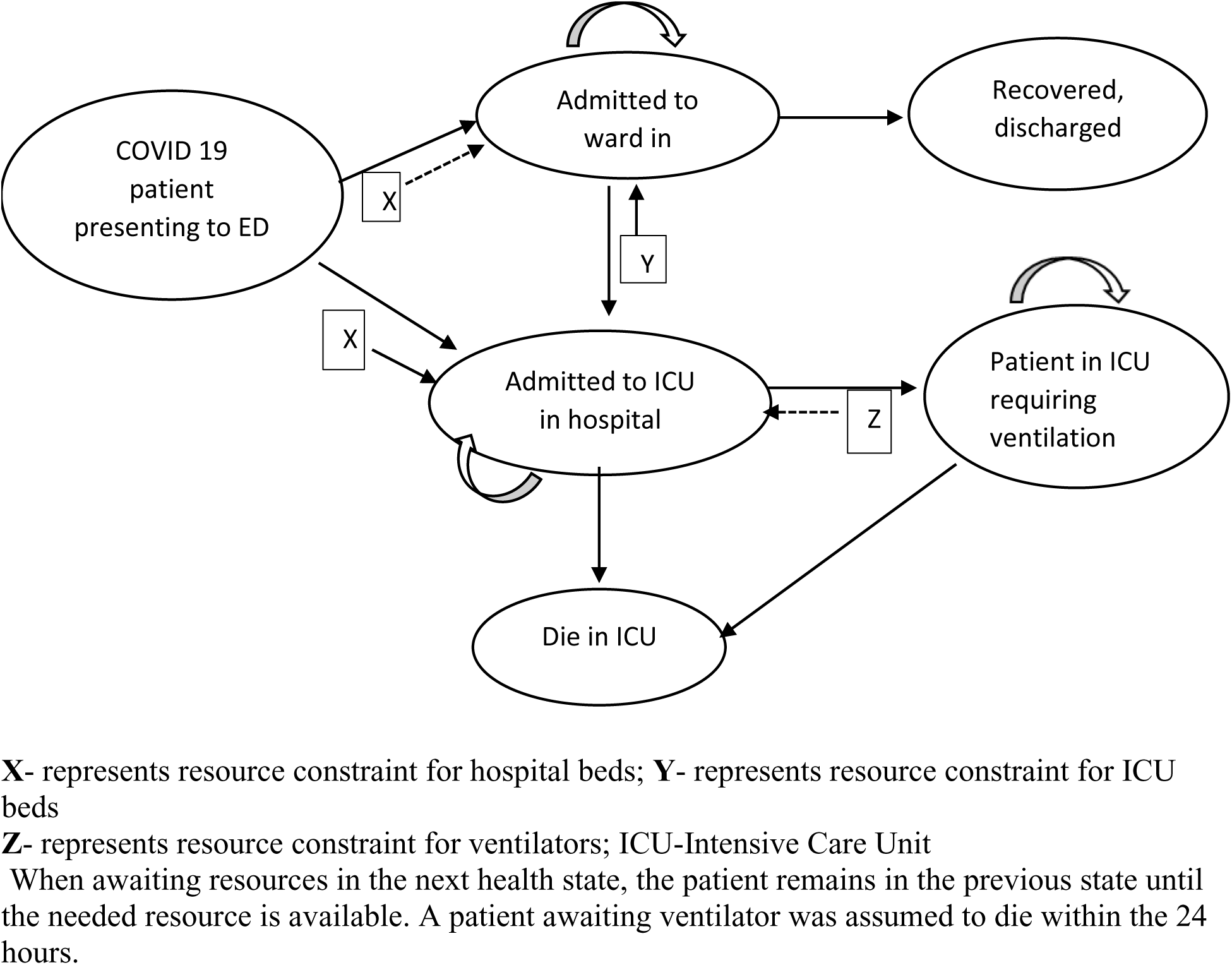
Model schematic.

**Figure 2:**
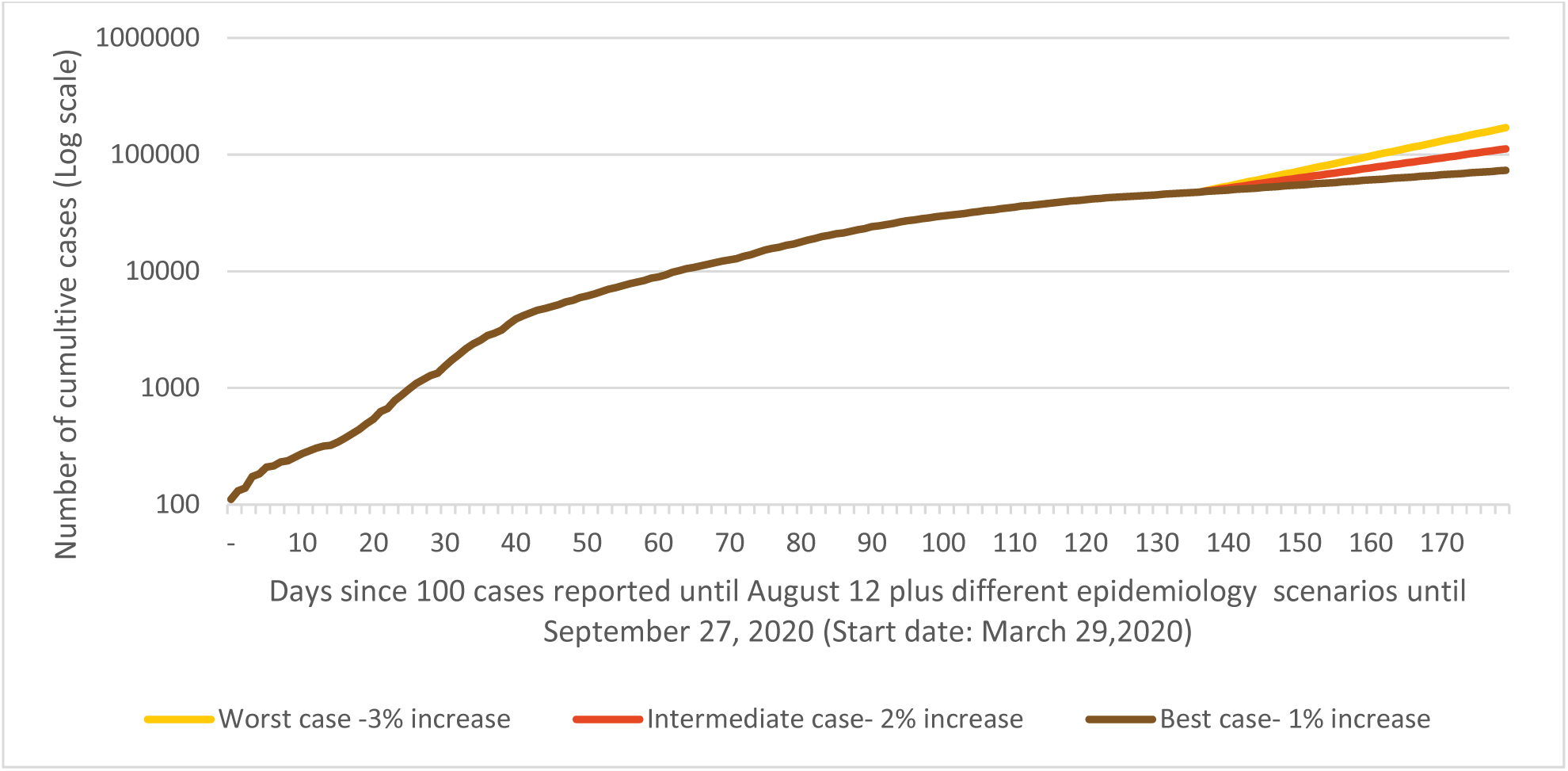
Cumulative cases of COVID-19 using different epidemic trajectories.

#### Resource capacity scenarios

We obtained data on the total number of ward beds, intensive care unit beds and ventilators before the pandemic from the published literature and press reports.^10,11,12^ Our data estimate for total ICU beds and ventilators were obtained from a current press report by Nigeria health officials assessing the available health resources in Nigeria. We modelled three resource scenarios: conservative, expanded and surplus. For the conservative scenario, we assumed that 50% of ventilated ICU beds (175/350), and 25% of hospital beds (10,000/40,000) are available to treat symptomatic patients based on recent press reports and a national survey conducted by a pharmaceutical company.^10,11,12^ In the expanded resource scenario, we added 200 ventilated ICU beds to the proportion available for treating COVID 19 ill-patients in conservative resource scenario and an additional 10,000 ward beds to the proportion of ward beds available in conservative resource scenario. The assumption was based on reduced clinical activity enabling re-allocation of privately-owned hospital resources, the recent increase in health resource supplies from donors, and the increase in government investment in healthcare services and resources since the start of the pandemic.^20^ For the surplus scenario, we assumed an additional 1,000 ventilated ICU beds to the proportion available for treating COVID 19 ill-patients in expanded resource scenario, and an extra 15,000 ward beds to the proportion of ward beds in expanded scenario resources based on potential funding from foreign donors, high-income countries and international health organizations.^21^ In the three hospital resource scenarios, we assumed adequate hospital staffing based on recent recall of retired health personnel to service by the Nigeria Federal government and the ongoing recruitment and training of volunteers across the country to boost staffing capacity.^22,23^

## RESULTS

For the best-case epidemic trajectory (1% growth rate), our study predicted 86,735 COVID-19 cases over 60 days (Figure 2), of whom 4,782 patients would require hospital admission to ward and ventilated ICU bed. In all resource scenarios, ward beds would remain available; ventilated ICU bed would be depleted after 5 days and 27 days in both conservative and expanded resource scenarios (Table 2).

**Table 2:** Estimated time to resource depletion by resource and epidemic trajectory scenarios.

|  |  | Estimated time to resource depletion (in days) |  | Number of avoidable deaths |
| --- | --- | --- | --- | --- |
|  |  | Ward beds | Ventilated ICU bed |  |
| ***Resource scenario: Conservative*** |  |  |  |  |
| Epidemic scenario | Best case (1% increase) | - | 5 | 901 |
|  | Intermediate (2% increase) | - | 5 | 1,703 |
|  | Worst-case (3% increase) | - | 5 | 2,729 |
| ***Resource scenario: Expanded*** |  |  |  |  |
| Epidemic scenario | Best case (1% increase) | - | 27 | 64 |
|  | Intermediate (2% increase) | - | 16 | 701 |
|  | Worst-case (3% increase) | - | 14 | 1,820 |
| ***Resource scenario: Surplus*** |  |  |  |  |
| Epidemic scenarios | Best case (1% increase) | - | - | - |
|  | Intermediate (2% increase) | - | - | - |
|  | Worst-case (3% increase) | - | 57 | 2 |

ICU, Intensive Care Unit

For the intermediate –epidemic trajectory (2% growth rate), we predicted 156,646 cumulative cases and 6,116 hospital admissions to ward and ventilated ICU bed over the 60-day time horizon. Although ward beds were not depleted in any of the resource scenarios, shortage of ventilated ICU bed would occur after 5 days in the conservative scenario. In the expanded scenario, ventilated ICU bed would be depleted after 16 days. In the surplus scenario, both resources would still be available after 60 days.

For the worst-case epidemic trajectory, we predicted 281,283 COVID-19 cases and 8,143 hospital admissions to ward and ventilated ICU beds over 60 days. In all three resource scenarios, ward beds would not be depleted. In the conservative resource scenario, ventilators would be unavailable after 5 days. However, by expanding ventilator supply as assumed in the surplus scenario, ventilators would be available until day 57 before a shortage is experienced.

Across all scenarios, sufficient resources to care for hospitalized COVID-19 patients would only be available in the best-case and intermediate epidemic trajectory in combination with the surplus resource scenario.

Assuming the conservative resource scenario, the best-case epidemic trajectory projects 901 preventable deaths over 60 days. In the best case and intermediate epidemic trajectories, assuming the surplus scenario, no death due to the unavailability of resources was predicted to occur over 60 days.

## DISCUSSION

We demonstrated that a substantial increase in healthcare resources is critical for Nigeria’s health system to care for COVID-19 patients and improve health outcomes, even at low infection growth rates. Maintaining aggressive public health measures are therefore needed to effectively reduce transmission, reduce the number of new cases and deaths. As of August 12, the public health measures implemented in Nigeria are moderate, and the average infection growth rate is approximately 5% (averaging from March 29 when 100 cases were reported to August 12, 2020). Since the implementation of public health measures, the average growth rate has further declined to 1-2 % within the last three months. While the Federal government acted promptly to initiate a lockdown in two of Nigeria’s most affected states on March 31, 2020, this decision was reversed in May due to economic pressures, despite the fluctuating low to the high daily number of cases.^24^ Our analysis shows that maintaining strong public health measures with vast-increase in resources is crucial to prevent hospital resource constraints and health system collapse in Nigeria.

Our study has limitations. Our estimated number of ward bed was based on a national survey by a pharmaceutical company published in 2007 and might not represent the country’s current capacity. While keeping with the current literature, we assumed that death would only occur in critically ill patients, which may underestimate mortality. Since the start of the pandemic, recruitment and training of new hospital staff have been ongoing, but actual data on the number of recruited staff was not available at the time of modelling, limiting our knowledge on the effect of vast expansion of hospital resources on staff capacity. Our study relies on reported cases to forecast future epidemic trajectories. The model does not account for underreporting of daily cases and long-time lag in case reports, which are common problems that occur in low resource settings due to limited testing capacity.^6^ Due to the unavailability of detailed COVID-19 data, some parameters included in the model were estimates obtained from a study on ARDS, a disease with similar clinical manifestations with COVID-19. Our healthcare resource utilization probabilities parameters (i.e. length of hospital stay and probability for need of ward, ICU and ventilator admissions and probability of death) were estimates from Canadian setting. While these parameter values arguably deviate from observed data in Africa, they are likely estimates for COVID-19 disease severity.

Our study has several strengths. We incorporated observed incidence data from Nigeria and other Africa countries with a range of epidemic trajectories to strengthen the validity of our epidemic predictions. The CORE model considers resource constraints within the health system and estimated deaths due to resource depletion. We stratified mortality between patients who receive adequate care in ICU and those who did not to estimate the number of patients who will likely die from an overwhelmed health system.

The COVID-19 epidemic trajectory is slower in Africa,^6^ with many countries within 5–10% average infection growth rates since 100 cases reported. Our epidemic trajectory and resource scenario predictions could apply to other low resource setting countries, especially in the Africa region. Our findings are supported by modelling studies that predict an expanding increase in the number of COVID-19 cases in Africa region^25^ and the need for strong government measures, abundancy of medical supplies and good personal protective behavior to mitigate the spread of COVID-19 in the region.^26^ Our resource scenarios estimates could also be used as a guide to inform pandemic preparedness planning and policy development.

## CONCLUSION

The epidemic trajectory of COVID-19 appears to have a low growth rate, and the implemented public health measures have helped to reduce community spread in Nigeria; however, the current hospital resources in Nigeria are still inadequate to manage the daily number of COVID-19 critically ill-patients. To mitigate the impact of COVID-19, implementing more aggressive public health measures is vital, and strategies to exponentially increase the number of health resources available in the country need to be put in place to prevent overwhelming the healthcare system.

## Data Availability

Study data derived from primary literature are publicly available to other researchers. Other data derived from health agencies are available on request directed to these agencies.

## Contributors

AO and BS designed the study, AO and AEJ acquired the data. AO, PP, SM, RX and BS conducted the data analysis. KB, YK, PP, SM, RX, DN and BS constructed the model. AO drafted the manuscript. All authors revised the manuscript critically and gave final approval of the version to be published. AO is the guarantor. The corresponding author attests that all listed authors meet authorship criteria and that no others meeting the criteria have been omitted.

## Transparency declaration

AO affirms that the manuscript is an honest, accurate, and transparent account of the study being reported; that no important aspects of the study have been omitted; and that any discrepancies from the study as originally planned (and, if relevant, registered) have been explained.

